# Standard Blood Laboratory as a Clinical Support Tool to Distinguish between SARS-CoV-2 Positive and Negative Patients

**DOI:** 10.1101/2020.10.23.20217844

**Authors:** Rainer Thell, Jascha Zimmermann, Marton Szell, Sabine Tomez, Philip Eisenburger, Moritz Haugk, Anna Kreil, Alexander Spiel, Amelie Blaschke, Anna Klicpera, Oskar Janata, Walter Krugluger, Christian Sebesta, Harald Herkner, Brenda Laky

## Abstract

**Background:** Coronavirus disease 2019 (COVID-19) caused by severe acute respiratory syndrome coronavirus 2 (SARS-CoV-2) is current pandemic disease. Acute polymerase-chain-reaction is the gold standard test for this disease, is not available everywhere. Standard blood laboratory parameters may have diagnostic potential.

**Methods:** We evaluated standard blood laboratory parameters of 655 COVID-19 patients suspected to be infected with SARS-CoV-2, who underwent PCR testing in one of five hospitals in Vienna, Austria. Additionally, clinical characteristics and 28-day outcome were obtained from medical records. We compared standard blood laboratory parameters, clinical characteristics, and outcomes between positive and negative PCR-tested patients and evaluated the ability of those parameters to distinguish between groups.

**Results:** Of the 590 study patients including 276 females and 314 males, aged between 20 and 100 years, 208 were tested positive by means of PCR. Patients with positive compared to negative PCR-tests had significantly lower levels of leukocytes, basophils, eosinophils, monocytes, and thrombocytes; while significantly higher levels were detected with hemoglobin, C-reactive-protein (CRP), neutrophil-to-lymphocyte ratio (NLR), activated-partial-thromboplastin-time (aPTT), creatine-kinase (CK), lactate-dehydrogenase (LDH), alanine-aminotransferase (ALT), aspartate-aminotransferase (AST), and lipase. Our multivariate model correctly classified 83.9% of cases with a sensitivity of 78.4%, specificity of 87.3%, positive predictive value of 79.5%, and negative predictive value of 86.6%. Decreasing leucocytes and eosinophils and increasing hemoglobin and CRP were significantly associated with an increased likelihood of being COVID-19 positive tested.

**Conclusions:** Our findings suggest that especially leucocytes, eosinophils, hemoglobin, and CRP are helpful to distinguish between COVID-19 positive and negative tested patients and that a certain blood pattern is able to predict PCR-results.

**Summary:** Decreasing leucocytes and eosinophils and increasing hemoglobin and CRP were significantly associated with an increased likelihood of being COVID-19 positive tested. Each single parameter showed either a high sensitivity (leucocytes, eosinophils, CRP, monocytes, thrombocytes) or specificity (NLR, CK, ALT, lipase), or a sensitivity and specificity around 60% (Hb, LDH, AST).

## INTRODUCTION

In December 2019, an uncommonly high incidence of pneumonia occurred in Wuhan province, China, caused by a previously unknown pathogen and showing an unusually high mortality rate, which showed to be a novel corona virus, SARS-CoV-2 (Severe Acute Respiratory Syndrome – Corona Virus 2).[1] It causes coronavirus disease (COVID-19) which has reached pandemic levels resulting in significant morbidity and mortality affecting all inhabited areas of the world with large numbers of patients. Hence, the World Health Organisation (WHO) declared a worldwide pandemic in March 2020.

Diagnostic steps for this disease currently are epidemiological contact history, clinical impression, chest radiography, standard blood laboratory, and antigen detection by means of real-time fluorescence PCR. PCR is the gold standard test for detection of SARS-CoV-2 infection.[2]

It soon became apparent that the available test capacities for PCR testing were far from sufficient, and a feverish search for alternative and simpler detection methods began. To date, point-of-care PCR testing is still not available everywhere. Testing that takes a long time can make up for significant additional efforts of organisation of patient cohorts in hospitals.

The clinical appearance of the disease is broadly reported.[3] Signs and symptoms may include fever and cough most commonly, dyspnea, rarely diarrhea, anosmia, or ageusia and others, at the beginning or during the course of the disease.[4-6] Various publications indicated that COVID-19 positive patients showed typical laboratory patterns,[7-9] although many of the earlier studies report on relatively small numbers of COVID-19 positive patients.

Zhang et al. included 95 cases with COVID-19-positive pneumonia.[7] They found significantly higher numbers of D-Dimer, C-reactive protein (CRP), and procalcitonin in patients to be admitted to intensive care levels. In a retrospective report of 138 admitted patients, lymphopenia occurred in 70%, a prolonged prothrombin time in 58%, and an elevated lactate dehydrogenase (LDH) resulted in 40% of patients.[10] In an Iranian cohort of 70 COVID-19 positive patients, Mardani et al. found significantly higher neutrophil count, CRP, LDH, aspartate aminotransferase (AST), alanine aminotransferase (ALT), and urea levels, as well as lowered white blood count and albumin levels.[11] A further retrospective trial of 99 patients showed decreased lymphocyte counts in 35%, decreased albumin in 98%, increased LDH in 75%, an increased interleukin-6 in 52%, and raised CRP in 86% of patients were reported.[3] Li et al. performed a trial with 989 patients, detecting a combination of eosinopenia and elevated CRP yielding a sensitivity of 67% and specificity of 78% for COVID-19.[12] In an additional cohort of 458 patients from Guan et al., leukopenia, lymphopenia, eosinopenia, and an increased CRP were detected.[4] In two trials, an increased NLR was described as an independent risk factor of mortality for COVID-19 patients.[13, 14]

Standard blood laboratory data are potentially of both, diagnostic and prognostic value.[15] On one hand, they can contribute to judge the pre-test probability of a COVID-19 diagnosis and thus, support the effective and efficacious organisational management of a patient in an emergency department. On the other hand, it is of potential value, if standard blood laboratory blood results can help to distinguish a potentially life-threatening course of a disease from a less critical status.

Therefore, the overall aim of this study was to evaluate standard blood laboratory parameters and clinical characteristics in a large number of COVID-19 suspicious patients, who underwent PCR testing. The specific aim was to determine whether standard blood laboratory parameters are able to identify positive COVID-19 patients from a large COVID-19 suspected cohort.

## MATERIALS AND METHODS

### Data Source

Data were obtained from an electronical data base of the Vienna Health Care Association (Wiener Gesundheitsverbund), which stores all medical records of patients treated in its hospitals in Vienna, Austria. Data extraction was exclusively performed by authorised employees of the emergency department at the Klinik Donaustadt, Vienna. The study protocol was approved both by the Ethics Commission of the City of Vienna (EK 20-122-VK) and the Ethics Commission of Sigmund Freud University Vienna (161/2020).

### Data Collection

Data were collected from female and male adult patients with suspected COVID-19 who underwent reverse transcriptase PCR testing via a nasopharyngeal swab performed at one of the five hospitals (Klinik Donaustadt, Klinik Floridsdorf, Klinik Hietzing, Klinik Landstrasse, and Klinik Ottakring) between February 27, 2020 and April 27, 2020. Based on PCR results, patients were divided into a COVID-19 positive or negative group. Patients without reported standard blood laboratory reports, medical history, or outcome documentation at day 28 after consultation were not included.

As standard blood laboratory testing was performed according to the clinical care needs of the patients, not all parameters were available for all patients. Parameters available from less than 20 patients in the positive or negative group were not analysed. Routine blood tests generally included full blood count, blood chemistry, electrolytes, liver function parameters, renal and myocardial function parameters as well as coagulation markers and markers of inflammation.

Patients’ gender (female/male), age at time of PCR testing (years), coexisting diseases and conditions (e.g. chronic diseases of the lung, liver, kidney; coronary artery disease, diabetes, and arterial hypertension); the clinical 28-day outcome including hospital admission and discharge as well as requirement of intensive care and ventilation, and death details were extracted from medical records.

### Statistical Analysis

Descriptive statistics was used to describe the characteristics of patients. The distribution of the data was determined by visual inspection of the histograms and the Kolmogorov Smirnov tests. Normally distributed data were calculated as mean value with standard deviation (SD), otherwise as median and interquartile range (IQR).

Continuous variables were compared between COVID-19 positive and negative patients with independent t-tests (parametric) or Mann-Whitney U-tests (non-parametric). Blood parameters were categorised according to normal reference ranges used in hospitals. However, since CRP ranges higher than normal (cut off: 0.5mg/dL) were detected in almost all patients (COVID-19 positive: 99.5% and negative: 98.2%) and thus, useless to discriminate between COVID-19 positive and negative patients, coordinates of the receiver operating characteristic (ROC) and the Youden index were used to determine another, sensitivity and specificity balanced, cut off value for CRP in our cohort. Chi-square or Fisher’s exact tests were applied to describe the relationship between proportions of categorical variables. Correlations between the continuous parameters were performed using Kendall’s Tau. Since neutrophils significantly correlated with leucocytes (0.859; p<0.001) and to reduce the number of predictive variables, we used the neutrophils-to-lymphocytes ratio (NLR) instead of neutrophils and lymphocytes. A cut-off for NLR was determined using coordinates of the ROC and the Youden index (sensitivity and specificity balanced). Hemoglobin was used instead of erythrocytes (0.761; p<0.001) and hematocrit (0.874; p<0.001) due to significant correlations. Procalcitonin and D-Dimer were also not included due to the small sample size. For further analysis between COVID-19 positive and negative patients, only significant different and clinical meaningful parameters were considered.

Univariate and multivariate binomial logistic regression analyses were used to construct prediction models using PCR results (COVID-19 positive/negative) as the dependent variable and significant patients’ characteristics and blood parameters as predictors (independent variables). Linearity of the continuous variables regarding the logit of the dependent variable was assessed using Box-Tidwell procedure with Bonferroni correction. None of the continuous variables violated the linearity assumption. None of the parameters showed multicollinearity.

Percentage accuracy in classification (PAC), sensitivity, specificity, positive predictive value (PPV), and negative predictive value (NPV) were estimated to assess models’ performance. Area under the ROC curves (AUC) were determined in order to assess the overall discriminatory ability of the model and of blood parameters to distinguish between COVID-19 positive and negative patients. Statistical significance was set at a p-value of <0.05 (two-sided). All data were analysed with SPSS software (IMP Statistics Version 25; SPSS Inc, Chicago, IL).

## RESULTS

### Characteristics of COVID-19 Suspected Patients

A total of 655 patients from the four hospitals underwent PCR testing between February 27, 2020 and April 27, 2020. Forty-five patients were not evaluated due to missing data. Another 17 patients who were tested within this time period with complete data sets were excluded since they were hospitalized more than one month before (n=16) or more than seven days after (n=1) PCR-testing. Patients (n=3) with both eosinophilia and acute malignant disease were excluded as well.

The median age of the 276 female (46.8%) and 314 male (53.2%) patients was 71 years (range, 20-100years; 60.7% of the patients were ≥65 years of age at time of PCR testing). No comorbidities were recorded in 69 (33.2%) and 84 (22.0%) COVID-19 positive and negative tested patients, respectively. COVID-19 negative tested patients had significantly more comorbidities than COVID-19 positive tested patients (median (IQR): 1 (1-3) vs. 1 (0-2); p <0.001). The most common comorbidity was pre-existing arterial hypertension (58.1%), followed by diabetes (25.4%), coronary heart disease (19.5%), chronic lung disease (16.9%), chronic kidney disease (15.8%), malignant diseases (13.6%), cerebrovascular accidents (7.5%), chronic liver disease (4.2%), and human immunodeficiency virus (HIV, 0.5%). A comparison between demographic characteristics and comorbidities between COVID-19 positive and negative tested patients showed no significant differences between the groups (Table 1).

**Table 1.** Comparison of Demographic Characteristics and 28-day Clinical Outcome between COVID-19 Positive and Negative tested Patients.

| Characteristic | COVID-19<br>Positive<br>(N=208) | COVID-19<br>Negative<br>(N=382) | P Value |
| --- | --- | --- | --- |
| Male – no./total no. (%) | 117 (56.3) | 197 (51.6) | 0.276* |
| Age |  |  |  |
| Median (IQR) - yr | 72 (57-79.75) | 70 (54-81) | 0.526<sup>†</sup> |
| Comorbidities – no./total no. (%) |  |  |  |
| Hypertension | 114 (54.8) | 229 (59.9) | 0.227* |
| Diabetes | 50 (24.0) | 100 (26.2) | 0.569* |
| Coronary heart disease* | 33 (15.9) | 82 (21.5) | 0.101* |
| Chronic lung disease<sup>†</sup> | 22 (10.6) | 78 (20.4) | 0.002* |
| Chronic kidney disease<sup>‡</sup> | 29 (13.9) | 64 (16.8) | 0.371* |
| Malignant tumor<sup>§</sup> | 12 (5.8) | 68 (17.8) | <0.001* |
| Cerebro vascular accident¶ | 13 (6.3) | 31 (8.1) | 0.410* |
| Chronic liver disease<sup>□</sup> | 2 (1.0) | 23 (6.0) | 0.004* |
| Human immunodeficiency virus | 0 | 3 (0.8) | 0.556<sup>‡</sup> |
| Clinical 28-day outcome – no. (%) |  |  |  |
| Not hospitalized/outpatient | 23 (11.1) | 33 (8.6) | <0.001* |
| Discharged after ≤28 days hospitalization<br>not requiring ICU support or ventilation | 97 (46.6) | 245 (64.1) |  |
| Discharged after ≤28 days hospitalization<br>requiring ICU support and/or ventilation | 4 (1.9) | 25 (6.5) |  |
| still in hospital after day 28 | 31 (14.9) | 44 (11.5) |  |
| Died before day 28 | 53 (25.5) | 35 (9.2) |  |

Abbreviation: COVID-19, coronavirus disease 2019; ICU, intensive care unit; IQR, interquartile range
\* Chi-square test; <sup>†</sup> Mann-Whitney U-test; <sup>‡</sup> Fischer's exact test

Similar numbers of outpatients tested positive (11.1%) or negative (8.6%; p=0.338). There was also no significant difference between COVID-19 positive compared to negative tested patients regarding the number of patients requiring ICU and/or ventilation (4.0% vs. 9.3%, respectively; p=0.091). Significantly more COVID-19 positive (25.5%) than negative (9.2%) tested patients died before day 28 (p<0.001). Main causes of death in COVID-19-positive patients were, in descending order, pneumonia (67.3%), followed by multi-organ failure (24.5%), acute cardiac failure (7.6%), and acute renal failure (1.9%).

### Comparison of Standard Blood Laboratory Parameters between Covid-19 Positive and Negative Patients

COVID-19 positive patients had significantly lower levels of leukocytes, NLR, basophils, eosinophils, monocytes, and thrombocytes; while significantly higher levels were detected with hemoglobin, CRP, aPTT, CK, LDH, ALT, AST, and lipase compared to COVID-19 negative patients. Similar levels were detected regarding procalcitonin, albumin, glucose, potassium, total bilirubin, GGT, creatinine, and BUN between the groups (Supplement Table 1). Parameters with significant differences are presented in Table 2.

**Table 2.** Comparison of Standard Blood Laboratory Parameters between COVID-19 Positive and Negative tested Patients.

| Parameters | N* | COVID-19<br>Positive | N* | COVID-19<br>Negative | P Value |
| --- | --- | --- | --- | --- | --- |
| *Blood count* |  |  |  |  |  |
| **Leucocytes (10<sup>9</sup>/L)** |  |  |  |  |  |
| Median (IQR) | 207 | 6.13 (4.80-8.08) | 378 | 9.06 (6.59-12.45) | <0.001<sup>†</sup> |
| Distribution – no./total no. (%) |  |  |  |  |  |
| Low +normal (≤10.0) | 179 | 86.5 | 221 | 58.5 | <0.001<sup>‡</sup> |
| High (>10.0) | 28 | 13.5 | 157 | 41.5 |  |
| **Neutrophil-to-lymphocyte ratio** |  |  |  |  |  |
| Median (IQR) | 534 | 5.00 (2.83-8.10) |  | 5.44 (3.30-9.25) | 0.032<sup>†</sup> |
| Distribution – no./total no. (%) |  |  |  |  |  |
| ≤2.33 | 36 | 19.8 | 27 | 7.7 | <0.001<sup>‡</sup> |
| >2.33 | 146 | 80.2 | 325 | 92.3 |  |
| **Basophils (10<sup>9</sup>/L)** |  |  |  |  |  |
| Median (IQR) | 182 | 0.02 (0.01-0.03) | 352 | 0.03 (0.02-0.04) | <0.001<sup>†</sup> |
| **Eosinophils (10<sup>9</sup>/L)** |  |  |  |  |  |
| Median (IQR) | 182 | 0.01 (0.00-0.05) | 352 | 0.10 (0.04-0.20) | <0.001<sup>†</sup> |
| Distribution – no./total no. (%) |  |  |  |  |  |
| Low (<0.1) | 155 | 85.2 | 176 | 50.0 | <0.001<sup>‡</sup> |
| Normal + high (≥0.1) | 27 | 14.8 | 176 | 50.0 |  |
| **Monocytes (10<sup>9</sup>/L)** |  |  |  |  |  |
| Median (IQR) | 182 | 0.47 (0.30-0.64) | 352 | 0.56 (0.40-0.77) | <0.001<sup>†</sup> |
| Distribution – no./total no. (%) |  |  |  |  |  |
| Low + normal (≤0.9) | 169 | 92.9 | 299 | 84.9 | 0.008<sup>‡</sup> |
| High (>0.9) | 13 | 7.1 | 53 | 15.1 |  |
| **Thrombocytes (10<sup>9</sup>/L)** |  |  |  |  |  |
| Median (IQR) | 207 | 201 (161-252) | 377 | 227 (169.5-299) | 0.002<sup>†</sup> |
| **Hemoglobin (g/dL)** |  |  |  |  |  |
| Median (IQR) | 207 | 13.50 (12.30-14.70) | 378 | 11.75 (10.28-13.33) | <0.001<sup>†</sup> |
| Distribution – no./total no. (%) |  |  |  |  |  |
| Low (f: <11.8; m: <13.5) | 67 | 32.4 | 234 | 61.9 | <0.001<sup>‡</sup> |
| Normal + high (f: ≥11.8; m: ≥13.5) | 140 | 67.6 | 144 | 38.1 |  |
| Inflammation |  |  |  |  |  |
| C-reactive protein (mg/dL) |  |  |  |  |  |
| Median (IQR) | 205 | 61.80 (25.45-129.50) | 381 | 33.60 (7.25-93.56) | <0.001<sup>†</sup> |
| Distribution – no./total no. (%) |  |  |  |  |  |
| <22 mg/dL | 44 | 21.5 | 161 | 42.3 | <0.001<sup>†</sup> |
| ≥22 mg/dL | 161 | 78.5 | 220 | 57.7 |  |
| Coagulation |  |  |  |  |  |
| Activated partial thromboplastin time (sec) |  |  |  |  |  |
| Median (IQR) | 141 | 29.90 (27.00-33.05) | 298 | 27.00 (24.00-31.00) | <0.001<sup>†</sup> |
| Heart function |  |  |  |  |  |
| Creatine Kinase (U/L) |  |  |  |  |  |
| Median (IQR) | 181 | 127.0 (63.0-243.5) | 336 | 102.5 (50.0-216.0) | 0.023<sup>†</sup> |
| Distribution – no./total no. (%) |  |  |  |  |  |
| Normal (≤190) | 113 | 62.4 | 246 | 73.2 | 0.011<sup>‡</sup> |
| High (>190) | 68 | 37.4 | 90 | 26.8 |  |
| Lactate dehydrogenase (U/L) |  |  |  |  |  |
| Median (IQR) | 157 | 285.0 (228.0-384.5) | 294 | 225.0 (189.0-297.75) | <0.001<sup>†</sup> |
| Distribution – no./total no. (%) |  |  |  |  |  |
| Normal (≤250) | 55 | 35.0 | 173 | 58.8 | <0.001<sup>‡</sup> |
| High (>250) | 102 | 65.0 | 121 | 41.2 |  |

Abbreviations: f, female; IQR, interquartile range; L, liter; m, male.
\* Number of parameters available. <sup>†</sup> Mann-Whitney U-test <sup>‡</sup> Chi-square test <sup>§</sup> Fisher's exact test

**Table 2 (continued). Comparison of Standard Blood Laboratory Parameters between COVID-19 Positive and Negative tested Patients.**
| Parameters | N* | COVID-19 | N* | COVID-19 | P Value |
| --- | --- | --- | --- | --- | --- |
|  |  | Positive |  | Negative |  |
| *Liver function* |  |  |  |  |  |
| Alanine aminotransferase (U/L) |  |  |  |  |  |
| Median (IQR) | 171 | 32.00 (21.00-53.00) | 346 | 25.00 (15.00-45.00) | 0.001<sup>†</sup> |
| Distribution – no./total no. (%) |  |  |  |  |  |
| Normal (≤45) | 111 | 64.9 | 261 | 75.4 | 0.012<sup>‡</sup> |
| High (>45) | 60 | 35.1 | 85 | 24.6 |  |
| Aspartate aminotransferase (U/L) |  |  |  |  |  |
| Median (IQR) | 124 | 47.00 (29.00-70.00) | 205 | 26.00 (20.00-52.00) | <0.001<sup>†</sup> |
| Distribution – no./total no. (%) |  |  |  |  |  |
| Normal (≤35) | 46 | 37.1 | 130 | 63.4 | <0.001<sup>‡</sup> |
| High (>35) | 78 | 62.9 | 75 | 36.6 |  |
| Lipase (U/L) |  |  |  |  |  |
| Median (IQR) | 151 | 39.00 (25.00-65.00) | 282 | 23.00 (14.00-41.00) | <0.001<sup>†</sup> |
| Distribution – no./total no. (%) |  |  |  |  |  |
| Normal (≤60.0) | 106 | 70.2 | 240 | 85.1 | <0.001<sup>‡</sup> |
| High (>60.0) | 45 | 29.8 | 42 | 14.9 |  |
| *Renal function* |  |  |  |  |  |
| Creatinine (mg/dL) |  |  |  |  |  |
| Distribution – no./total no. (%) |  |  |  |  |  |
| Low (<0.5) | 0 | 0 | 12 | 3.2 | 0.009<sup>‡</sup> |
| Normal + high (≥0.5) | 208 | 100.0 | 365 | 96.8 |  |

Abbreviations: f, female; IQR, interquartile range; L, liter; m, male.
\* Number of parameters available. <sup>†</sup> Mann-Whitney U-test <sup>‡</sup> Chi-square test <sup>§</sup> Fisher's exact test

After dichotomizing blood laboratory parameters, similar significances were detected (Table 2); while binary comparisons of aPTT (normal+high (≥21sec): 97.9% positive vs. 97.6% negative) and total bilirubin (normal+high (≥0.3mg/dL): 93.6% positive vs. 89.6% negative) showed no significant differences between positive and negative tested patients (p=0.885 and p=0.132, respectively).

### Univariate and Multivariate Analyses of Standard Blood Laboratory Parameters

Univariate logistic regression analysis with COVID-19 positive tested patients as the dependent variable revealed that the majority of blood parameters including the dichotomized levels were associated with a positive COVID-19 diagnosis (Table 3). Comorbidities due to their low discrimination ability and parameters without significant associations were not included in further multivariate analyses (Supplement Table 2).

**Table 3.** Uni- and Multivariate Analyses of Standard Blood Laboratory Parameters with COVID-19 Positive tested Patients as the Dependent Variable.

| Parameters | Univariate<br>OR (95% CI) | P Value | Multivariate<br>OR (95% CI) | P Value |
| --- | --- | --- | --- | --- |
| *Blood count* |  |  |  |  |
| Leukocytes (10<sup>9</sup>/L) | 0.81 (0.77-0.86) | <0.001 | 0.69 (0.58-0.83) | <0.001 |
| Leukocytes (ref: >10.0) | 1.00 |  |  |  |
| leukopenia+normal (≤10.0) | 4.54 (2.90-7.11) | <0.001 | - | - |
| Neutrophil-to-lymphocyte ratio | 1.00 (0.98-1.02) | 0.745 | - | - |
| Neutrophil-to-lymphocyte ratio (ref: >2.33) | 1.00 |  |  |  |
| ≤2.33 | 2.97 (1.74-5.07) | <0.001 | 1.72 (0.45-6.58) | 0.428 |
| Basophils (10<sup>9</sup>/L) | 9.55*10<sup>-18</sup> | <0.001 | 0.00 | 0.133 |
|  | (6.02*10<sup>-23</sup>-1.51*10<sup>-12</sup>) |  | (0.00-1334.76) |  |
| Eosinophils (10<sup>9</sup>/L) | 0.00005 (0.000003-0.001) | <0.001 | 0.00 (0.00-0.97) | 0.011 |
| Eosinophils (ref: ≥0.1) | 1.00 |  |  |  |
| eosinopenia (<0.1) | 5.74 (3.63-9.09) | <0.001 | - | - |
| Monocytes (10<sup>9</sup>/L) | 0.26 (0.13-0.51) | <0.001 | 0.93 (0.12-7.06) | 0.942 |
| Monocytes (ref: >0.9) | 1.00 |  |  |  |
| monocytopenia+normal (≤0.9) | 2.30(1.22-4.35) | 0.010 | - | - |
| Thrombocytes (10<sup>9</sup>/L) | 0.998 (0.997-1.00) | 0.013 | 1.00 (1.00-1.01) | 0.272 |
| Thrombocytes (>370) | 1.00 |  |  |  |
| thrombopenia+normal (≤370) | 2.00 (1.10-3.66) | 0.024 | - | - |
| Hemoglobin (g/dL) | 1.39 (1.27-1.52) | <0.001 | 1.63 (1.29-2.07) | <0.001 |
| Hemoglobin (ref: f: <11.8; m: <13.5) | 1.00 |  |  |  |
| normal+high (f: ≥11.8; m: ≥13.5) | 3.40 (2.38-4.86) | <0.001 | - | - |
| *Inflammation* |  |  |  |  |
| C-reactive protein (U/L) | 1.004 (1.002-1.006) | <0.001 | 1.01 (1.00-1.02) | 0.025 |
| C-reactive protein (ref: <22 mg/dL) | 1.00 |  |  |  |
| ≥22 mg/dL | 2.68 (1.81-3.96) | <0.001 | - | - |
| *Coagulation* |  |  |  |  |
| Activated partial thromboplastin time (sec) | 1.04 (1.01-1.08) | 0.005 | 0.99 (0.92-1.06) | 0.683 |
| *Heart function* |  |  |  |  |
| Creatine Kinase (U/L) | 1.00004 (0.9999-1.0002) | 0.675 | - | - |
| Creatine kinase (ref: ≤190) | 1.00 |  |  |  |
| high (>190) | 1.65 (1.12-2.42) | 0.011 | 0.84 (0.28-2.59) | 0.767 |
| Lactate dehydrogenase (U/L) | 1.0005 (0.9996-1.001) | 0.274 | - | - |
| Lactate dehydrogenase (ref: ≤250) | 1.00 |  |  |  |
| high (>250) | 2.65 (1.77-3.96) | <0.001 | 2.36 (0.71-7.83) | 0.159 |
| *Liver function* |  |  |  |  |
| Alanine aminotransferase (U/L) | 0.999 (0.997-1.002) | 0.519 | - | - |
| Alanine aminotransferase (ref: ≤45) | 1.00 |  |  |  |
| high (>45) | 1.66 (1.12-2.47) | 0.013 | 1.65 (0.55-4.96) | 0.375 |
| Aspartate aminotransferase (U/L) | 1.000 (0.998-1.001) | 0.653 | - | - |
| Aspartate aminotransferase (ref: ≤35) | 1.00 |  |  |  |
| high (>35) | 2.94 (1.85-4.67) | <0.001 | 1.39 (0.44-4.38) | 0.573 |
| Lipase (U/L) | 1.002 (0.999-1.005) | 0.137 | - | - |
| Lipase (ref: ≤60) | 1.00 |  |  |  |
| high (>60) | 2.43 (1.50-3.92) | <0.001 | 1.44 (0.42-4.97) | 0.565 |
| *Renal function* |  |  |  |  |
| Creatinine (ref: 0.5-1.2mg/dL) | 1.00 |  | - | - |
| low (<0.5) | 0.00 | 0.999 |  |  |
| high (>1.2) | 1.26 (0.87-1.82) | 0.214 |  |  |

Abbreviation: CI, confidence interval; OR, odds ratio.

The multivariate logistic regression model was statistically significant (Chi-square (14) = 119.2; p<0.001) and explained 62.8% (Nagelkerke *R*^2^) of the variance in COVID-19 positive tested patients and correctly classified 83.9% of cases. Sensitivity was 78.4%, specificity was 87.3%, positive predictive value was 79.5%, and negative predictive value was 86.6%. Of the 14 predictors, four were statistically significant including leucocytes, eosinophils, hemoglobin, and CRP (Table 3). Decreasing leucocytes and eosinophils as well as increasing hemoglobin and CRP were associated with an increased likelihood of being COVID-19 positive tested. AUC, as a measure of the overall discriminatory ability of the model, and the model’s best blood parameters combined are presented as ROC curves in Figure 1.

**Figure 1.**
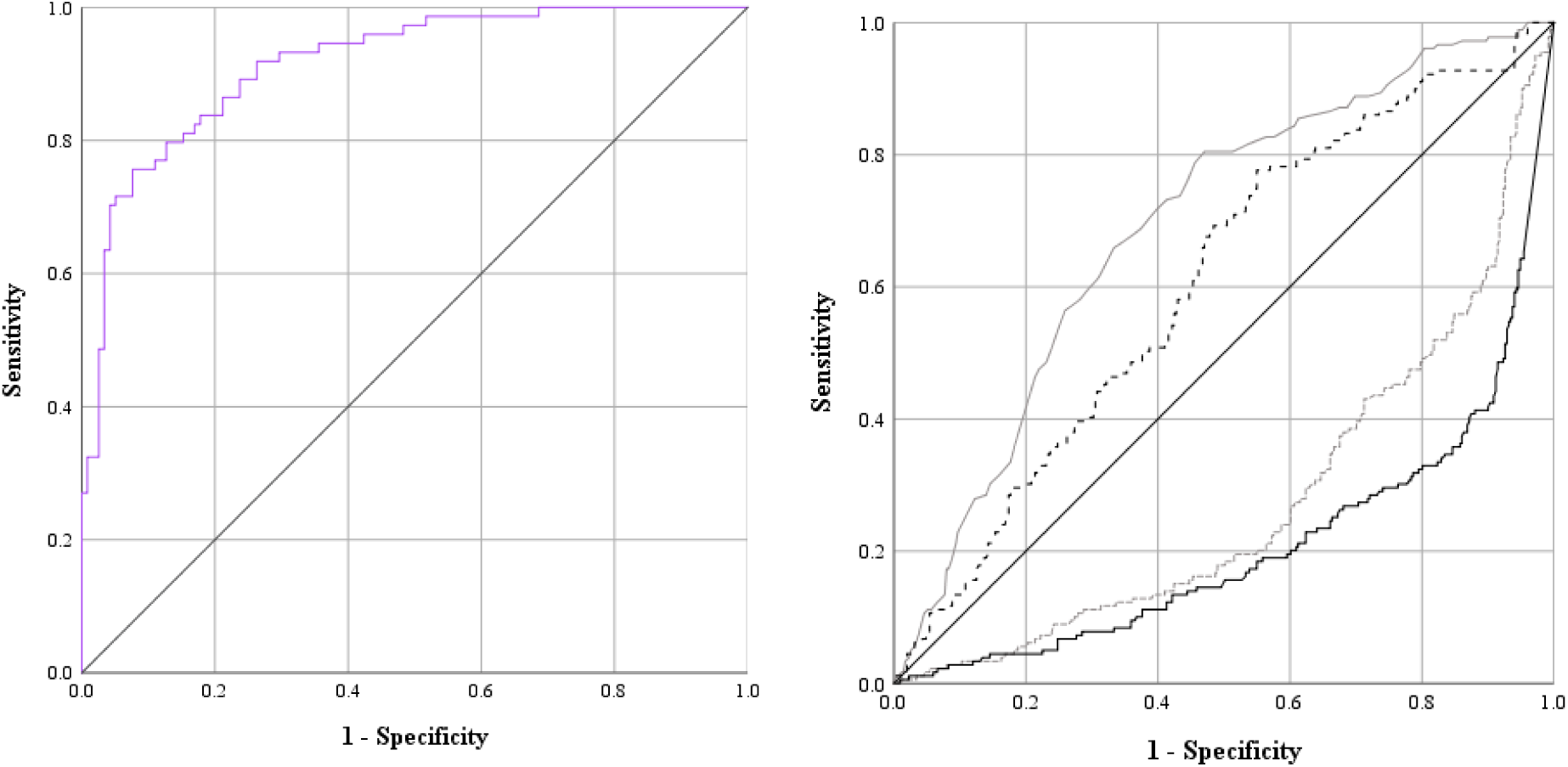
Receiver operating characteristic (ROC) curve of (A) the model with and area under the ROC curve (AUC) of 0.915 (95% confidence intervals (CI), 0.876 to 0.955) and (B) the combined blood parameters leucocytes (doted grey line; AUC=0.278, 95%CI, 0.232 to 0.324), eosinophils (straight black line; AUC=0.208, 95%CI, 0.165 to 0.250), hemoglobin (straight grey line; AUC=0.693, 95%CI, 0.646 to 0.739), and CRP (doted black line; AUC=0.605, 95%CI, 0.555 to 0.655).

### Diagnostic Performance of Single Standard Blood Laboratory Parameters

Diagnostic performance of those four dichotomized blood parameters, which were significant at the multivariate logistic regression as well as those, which were significant at the univariate analysis, were evaluated regarding their ability as single parameter to distinguish between COVID-19 positive from negative tested patients (Table 4). Each single parameter showed either a high sensitivity (leucocytes, eosinophils, CRP, monocytes, platelets), a high specificity (NLR, CK, ALT, lipase), or a sensitivity and specificity around 60% (hemoglobin, LDH, AST).

**Table 4.** Diagnostic Performance of Single Standard Blood Laboratory Parameters to Distinguish Between COVID-19 Positive from Negative Tested Patients.

| Parameters | Sensitivity<br>(%) | Specificity<br>(%) | PPV<br>(%) | NPV<br>(%) | AUC<br>(95% CI) | P<br>value |
| --- | --- | --- | --- | --- | --- | --- |
| Leukocytes<br>($\leq 10.0 \times 10^9/L$) | 86.5 | 41.5 | 44.8 | 84.9 | 0.640 (0.595-0.685) | <0.001 |
| NLR<br>($\leq 2.33$) | 19.8 | 92.3 | 57.1 | 69.0 | 0.561 (0.508-0.613) | 0.022 |
| Eosinophils<br>($< 0.10 \times 10^9/L$) | 85.2 | 50.0 | 46.8 | 86.7 | 0.676 (0.630-0.722) | <0.001 |
| Monocytes<br>($\leq 0.9 \times 10^9/L$) | 92.9 | 15.1 | 36.1 | 80.3 | 0.540 (0.489-0.590) | 0.134 |
| Thrombocytes<br>($\leq 370 \times 10^9/L$) | 92.8 | 13.5 | 37.1 | 77.3 | 0.209 (0.483-0.580) | 0.209 |
| Hemoglobin<br>(f: $\geq 11.8$; m: $\geq 13.5$ g/dL) | 67.6 | 61.9 | 49.3 | 77.7 | 0.648 (0.601-0.694) | <0.001 |
| CRP<br>($\geq 22$ mg/dL) | 78.5 | 42.3 | 42.3 | 78.5 | 0.604 (0.557-0.651) | <0.001 |
| CK<br>($> 190$ U/L) | 37.6 | 73.2 | 43.0 | 68.5 | 0.554 (0.501-0.606) | 0.043 |
| LDH<br>($> 250$ U/L) | 65.0 | 58.8 | 45.7 | 75.9 | 0.619 (0.565-0.673) | <0.001 |
| ALT<br>($> 45$ U/L) | 35.1 | 75.4 | 41.4 | 70.2 | 0.553 (0.499-0.606) | 0.051 |
| AST<br>($> 35$ U/L) | 62.9 | 63.4 | 51.0 | 73.9 | 0.632 (0.569-0.694) | <0.001 |
| Lipase<br>($> 60$ U/L) | 29.8 | 85.1 | 51.7 | 69.4 | 0.575 (0.517-0.632) | 0.011 |

Abbreviations: ALT, alanine aminotransferase; AST, aspartate aminotransferase; CK, creatine kinase; CRP, C-reactive protein; LDH, lactate dehydrogenase; NLR, Neutrophil-to-lymphocyte ratio.

## DISCUSSION

In this trial, we showed that the likelihood of a SARS-CoV-2 infection can be enforced through standard laboratory blood findings to a high degree. Several studies including meta-analyses recently focused on prediction of the severity of the disease derived from blood results.[8, 9, 16] Our consideration to find a model to predict the diagnosis of SARS-CoV-2 infection with standard blood parameters has been less studied.

To our knowledge, only three other trials comparing standard blood parameters between positive and negative cases are published to date.[11, 12, 17] Similar to those studies, our study showed that leucopenia, eosinopenia, elevated hemoglobin, and CRP were detected to be among the best standard laboratory parameters to distinguish between COVID-19 positive from negative tested patients. Accordingly, similar patterns have been detected in positive COVID-19 patients with a severe compared to a mild form of the disease.[8, 9, 16] The major differences of the three studies, which compared COVID-19 positive and negative patients, opposed to the reviews, which reported only on positive tested patients, were documented regarding leucocytes, neutrophils, and hemoglobin (Table 5).

**Table 5.**
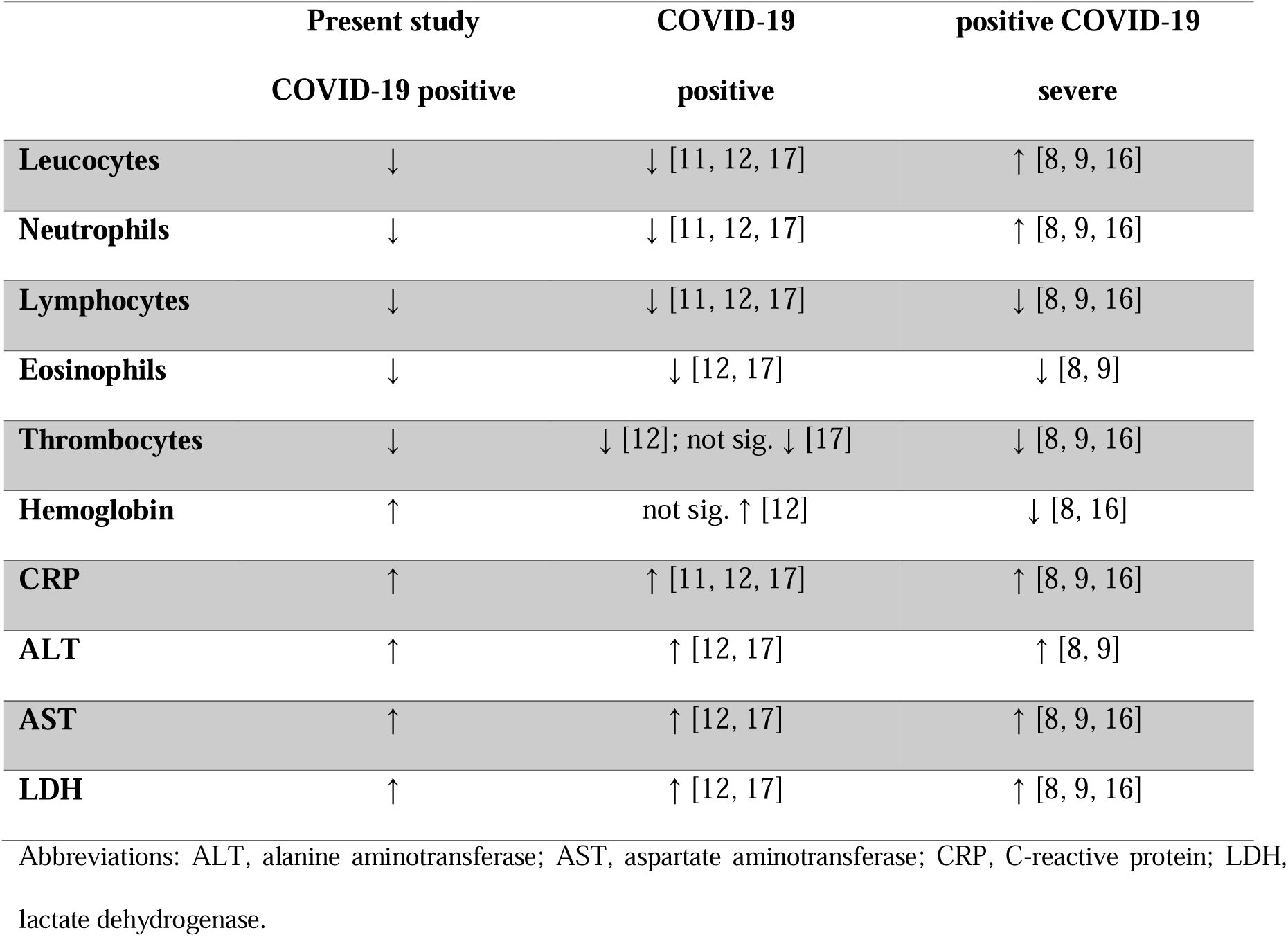
Blood parameter patterns of COVID-19 positive tested patients of various studies.

|  | **Present study** | **COVID-19** | **positive COVID-19** |
| --- | --- | --- | --- |
|  | **COVID-19 positive** | **positive** | **severe** |
| **Leucocytes** | ↓ | ↓ [11, 12, 17] | ↑ [8, 9, 16] |
| **Neutrophils** | ↓ | ↓ [11, 12, 17] | ↑ [8, 9, 16] |
| **Lymphocytes** | ↓ | ↓ [11, 12, 17] | ↓ [8, 9, 16] |
| **Eosinophils** | ↓ | ↓ [12, 17] | ↓ [8, 9] |
| **Thrombocytes** | ↓ | ↓ [12]; not sig. ↓ [17] | ↓ [8, 9, 16] |
| **Hemoglobin** | ↑ | not sig. ↑ [12] | ↓ [8, 16] |
| **CRP** | ↑ | ↑ [11, 12, 17] | ↑ [8, 9, 16] |
| **ALT** | ↑ | ↑ [12, 17] | ↑ [8, 9] |
| **AST** | ↑ | ↑ [12, 17] | ↑ [8, 9, 16] |
| **LDH** | ↑ | ↑ [12, 17] | ↑ [8, 9, 16] |

Abbreviations: ALT, alanine aminotransferase; AST, aspartate aminotransferase; CRP, C-reactive protein; LDH, lactate dehydrogenase.

In conformity with other publications [11, 12, 17], leucocytes were lower in COVID-19 positive than negative patients at the time of PCR testing, and so were neutrophils and lymphocytes; while severe compared to mild forms of COVID-19 tend to have higher leucocytes and neutrophils. As opposed to our findings, which showed a low ability of NLR to discriminate between positive and negative (AUC=0.561) and basically no contribution in the multivariate analysis, a raised NLR, which evolved from a raised neutrophil count as well as a lowered lymphocyte count, was already shown previously to be a prognostic value for endotracheal intubation and mortality predictor.[13, 14] A cut-off of 4.94 was used in the publication by Tatum et al. [14]; above this value, the risk of being artificially ventilated or to die was increased. Notably, 89% of those patients were African Americans. A lower cut-off (2.33) was established in our study, which might be because only 15% of patients had a neutrophil count higher than 7.7×10^9^/L. We are however unaware of any study using NLR as a pure discriminator between positive and negative COVID-19 diagnosis.

However, severity of illness appears to be less important regarding the other parameters, especially regarding eosinophils and CRP (Table 5). Like in other publications [12, 17], our data also showed that eosinopenia was one of the significant predictive biomarkers for COVID-19 with a sensitivity of 85% and a specificity of 50%.

Li et al. [12] and our study showed an increased hemoglobin in COVID-19 positive patients, which is not in accordance with a lowered hemoglobin in patients with severe COVID-19 disease reported by two meta-analyses.[8, 16] In our data, the median hemoglobin was 13.5 g/dL, which did not much differ from Li’s data.[12] In several other trials assessing the severity of disease and blood patterns, hemoglobin was shown to be below normal ranges.[8, 9, 16] It can only be hypothesized why our cohort presented with a comparably high level of hemoglobin. Possibly, a degree of dehydration played a role at the time of presentation in the emergency department. Indeed, an average temperature of 38.0±0.9°C on presentation in 99 COVID-19 positive and 37.1±1.4°C in 103 COVID-19 negative patients, which was a significant difference, was detected in a subgroup analysis of 202 of our patients.

Not surprisingly, CRP was significantly elevated in all studies.[11, 12, 17] In our patients, we set a new cut-off using the Youden index of 22 mg/dL, since the vast majority of patients had increased CRP values.

Similar blood patterns were also detected regarding ALT, AST, and LDH.[8, 9, 12, 16, 17] Brinati et al. included 279 patients and developed a score for SARS-Cov-2 detection with an accuracy between 82% and 86%, and sensitivity between 92% and 95%.[18] Applying our data including age, gender, leucocytes, neutrophils, lymphocytes, monocytes, eosinophils, basophils, thrombocytes, CRP, AST, ALT, GGT, and LDH to Brinati’s tool, a quite high AUC (AUC=0.709, 95%CI 0.646-0.771; p<0.001), sensitivity (70.4%), specificity (71.3%), and NPV (79.9%), but less promising PPV (59.8 %) could be obtained. However, our model including 14 standard laboratory blood parameters reached better diagnostic performances in all areas (AUC=0.915, 95%CI 0.876-0.955); sensitivity (78.4%); specificity (87.3%), PPV (79.5%), and NNP (86.6%)), although, the most prominent parameters were leucocytes, eosinophils, hemoglobin, and CRP.

The following limitations of the study should be noted. The retrospective design with missing blood parameters and no validation cohort are amongst the major limiting factors and thus, our selected model may be over-fitted. Additionally, with the single time point evaluation, we were not able to retrieve information regarding progression of the disease. Furthermore, cytokines, especially interleukin-6, were not routinely measured, which may be better predictors, especially regarding the so-called, COVID-19 cytokine storm’, to elucidate COVID-19 positive from negative patients. Another fact to consider is the heterogeneity of underlying diseases, which may also contribute to variations in our findings. On the other hand, such a heterogeneity may reflect reality during a pandemic situation best. Eventually, all test quality crucially depends on the quality of the manual specimen acquisition.[19, 20] PCR results tend to be more positive in patients with an increased viral load and with a shorter duration of the disease.[21]

## Conclusions

Generally, as laboratory equipment supply develops, more PCR point-of-care diagnostics become available. It is nonetheless doubtful that - neither in the near, nor in the far future – PCR will entirely replace standard laboratory testing. Therefore, the question of a blood laboratory pattern, as specific as possible for COVID-19, remains relevant. Our multivariate model including 14 standard blood parameters correctly classified 84% of cases with a sensitivity of 78% and a specificity of 87%, and therefore could be a useful model to facilitate rapid triage of potential COVID-19 patients.

## Data Availability

All data acquired can be reviewed by any editor as wished.

## Acknowledgment

All authors declare that they have no conflict of interest.

