## Supplement for "Standard Blood Laboratory as a Clinical Support Tool to Distinguish between SARS-CoV-2 Positive and Negative Patients"

**Supplement Table 1. Comparison of Standard Blood Laboratory Parameters between COVID-19 Positive and Negative tested Patients.**

| Parameters | N* | COVID-19 TOTAL | N* | COVID-19 Positive | N* | COVID-19 Negative | P value | Pattern |
| --- | --- | --- | --- | --- | --- | --- | --- | --- |
| *Blood count* |  |  |  |  |  |  |  |  |
| White blood cell count or Leucocytes (10^9^/L) |  |  |  |  |  |  |  |  |
| Median (IQR) | 585 | 7.90 (5.70 to 11.10) | 207 | 6.13 (4.80 to 8.08) | 378 | 9.06 (6.59 to 12.45) | <0.001^†^ | ↓ |
| Distribution – no./total no. (%) |  |  |  |  |  |  |  |  |
| Low (<4.0) | 44 | 7.5 | 24 | 11.6 | 20 | 5.3 | <0.001^‡^ |  |
| Normal (4.0-10.0) | 356 | 60.9 | 155 | 74.9 | 201 | 53.2 |  | ⊥ (not ↑) |
| High (>10.0) | 185 | 31.6 | 28 | 13.5 | 157 | 41.5 |  |  |
| Neutrophils (10^9^/L) |  |  |  |  |  |  |  |  |
| Median (IQR) | 534 | 5.79 (4.06 to 8.46) | 182 | 4.45 (3.12 to 6.62) | 352 | 6.66 (4.67 to 9.78) | <0.001^†^ | ↓ |
| Distribution – no./total no. (%) |  |  |  |  |  |  |  |  |
| Low (<1.5) | 4 | 0.7 | 4 | 2.2 | 0 | 0 | <0.001^‡^ |  |
| Normal (1.5-7.7) | 366 | 68.5 | 150 | 82.4 | 216 | 61.4 |  | ⊥ (not ↑) |
| High (>7.7) | 164 | 30.7 | 28 | 15.4 | 136 | 38.6 |  |  |
| Basophils (10^9^/L) |  |  |  |  |  |  |  |  |
| Median (IQR) | 534 | 0.02 (0.01 to 0.04) | 182 | 0.02 (0.01 to 0.03) | 352 | 0.03 (0.02 to 0.04) | <0.001^†^ | ↓ |
| Distribution – no./total no. (%) |  |  |  |  |  |  |  |  |
| Normal (0.0-0.2) | 532 | 99.6 | 182 | 100 | 350 | 99.4 | 0.550^§^ | ⊥ (=) |
| High (>0.2) | 2 | 0.4 | 0 | 0 | 2 | 0.6 |  |  |
| Eosinophils (10^9^/L) |  |  |  |  |  |  |  |  |
| Median (IQR) | 534 | 0.06 (0.01 to 0.16) | 182 | 0.01 (0.00 to 0.05) | 352 | 0.10 (0.04 to 0.20) | <0.001^†^ | ↓ |
| Distribution – no./total no. (%) |  |  |  |  |  |  |  |  |
| Low (<0.1) | 331 | 62.0 | 155 | 85.2 | 176 | 50.0 | <0.001^‡^ |  |
| Normal (0.1-0.5) | 195 | 36.5 | 26 | 14.3 | 169 | 48.0 |  | ↓ |
| High (>0.5) | 8 | 1.5 | 1 | 0.5 | 7 | 2.0 |  |  |
| Lymphocytes (10^9^/L) |  |  |  |  |  |  |  |  |
| Median (IQR) | 534 | 1.10 (0.75 to 1.55) | 182 | 0.96 (0.65 to 1.36) | 352 | 1.21 (0.84 to 1.70) | <0.001^†^ | ↓ |
| Distribution – no./total no. (%) |  |  |  |  |  |  |  |  |
| Low (<1.5) | 382 | 71.5 | 150 | 82.4 | 232 | 65.9 | <0.001^‡^ |  |
| Normal (1.5-4.5) | 151 | 28.3 | 31 | 17.0 | 120 | 34.1 |  | ↓ (less =) |
| High (>4.5) | 1 | 0.2 | 1 | 0.6 | 0 | 0 |  |  |
| Neutrophil-to-lymphocyte ratio |  |  |  |  |  |  |  |  |
| Median (IQR) | 534 | 5.29 (3.16-8.90) | 534 | 5.00 (2.83-8.10) |  | 5.44 (3.30-9.25) | 0.032^†^ | ↓ |
| Distribution – no./total no. (%) |  |  |  |  |  |  |  |  |
| ≤2.33 | 63 | 11.8 | 36 | 19.8 | 27 | 7.7 | <0.001^‡^ | more ↓ |
| >2.33 | 471 | 88.2 | 146 | 80.2 | 325 | 92.3 |  |  |
| Monocytes (10^9^/L) |  |  |  |  |  |  |  |  |
| Median (IQR) | 534 | 0.53 (0.36 to 0.74) | 182 | 0.47 (0.30 to 0.64) | 352 | 0.56 (0.40 to 0.77) | <0.001^†^ | ↓ |
| Distribution – no./total no. (%) |  |  |  |  |  |  |  |  |
| Low (<0.1) | 3 | 0.6 | 0 | 0 | 3 | 0.9 | 0.013^‡^ |  |
| Normal (0.1-0.9) | 465 | 87.0 | 169 | 92.9 | 296 | 84.0 |  | ⊥ (less ↓) |
| High (>0.9) | 66 | 12.4 | 13 | 7.1 | 53 | 15.1 |  |  |

**Supplement Table 1 (cont.). Comparison of Standard Blood Laboratory Parameters between COVID-19 Positive and Negative tested Patients.**

| Parameters | N* | COVID-19  TOTAL | N* | COVID-19  Positive | N* | COVID-19  Negative | P value | Pattern |
| --- | --- | --- | --- | --- | --- | --- | --- | --- |
| *Blood count (cont.)* |  |  |  |  |  |  |  |  |
| Platelet count or Thrombocytes (10^9^/L) |  |  |  |  |  |  |  |  |
| Median (IQR) | 584 | 217.00 (166.25 to 282.75) | 207 | 201.00 (161.00 to 252.00) | 377 | 227.00 (169.50 to 299.00) | 0.002^†^ | ↓ |
| Distribution – no./total no. (%) |  |  |  |  |  |  |  |  |
| Low (<150) | 104 | 17.8 | 35 | 16.9 | 69 | 18.3 | 0.052^‡^ |  |
| Normal (150-370) | 414 | 70.9 | 157 | 75.9 | 257 | 68.2 |  | ⊥ (=) |
| High (>370) | 66 | 11.3 | 15 | 7.2 | 51 | 13.5 |  |  |
| Red blood cell count or Erythrocytes (10^12^/L) |  |  |  |  |  |  |  |  |
| Median (IQR) | 585 | 4.20 (3.60 to 4.71) | 207 | 4.60 (4.11 to 5.00) | 378 | 3.90 (3.40 to 4.50) | <0.001^†^ | ↑ |
| Distribution – no./total no. (%) |  |  |  |  |  |  |  |  |
| Low (<4.3) | 310 | 53.0 | 67 | 32.4 | 243 | 64.3 | <0.001^‡^ |  |
| Normal (4.3-5.7) | 270 | 46.2 | 136 | 65.7 | 134 | 35.4 |  | ⊥ (not ↓) |
| High (>5.7) | 5 | 0.8 | 4 | 1.9 | 1 | 0.3 |  |  |
| Hemoglobin (g/dL) |  |  |  |  |  |  |  |  |
| Median (IQR) | 585 | 12.50 (10.70 to 14.10) | 207 | 13.50 (12.30 to 14.70) | 378 | 11.75 (10.28 to 13.33) | <0.001^†^ | ↑ |
| Distribution – no./total no. (%) |  |  |  |  |  |  |  |  |
| Low (f: <11.8; m: <13.5) | 301 | 51.5 | 67 | 32.4 | 234 | 61.9 | <0.001^‡^ |  |
| Normal (f: 11.8-15.8; m: 13.5-17.2) | 279 | 47.7 | 138 | 66.6 | 141 | 37.3 |  | ⊥ (not ↓) |
| High (f: 15.8; m: >17.2) | 5 | 0.8 | 2 | 1.0 | 3 | 0.8 |  |  |
| Hematocrit (%) |  |  |  |  |  |  |  |  |
| Median (IQR) | 585 | 37.20 (32.40 to 41.55) | 207 | 39.70 (36.20 to 42.80) | 378 | 35.70 (31.05 to 40.13) | <0.001^†^ | ↑ |
| Distribution – no./total no. (%) |  |  |  |  |  |  |  |  |
| Low (f: <38.0; m: <39.5) | 354 | 60.5 | 90 | 43.5 | 264 | 69.9 | <0.001^‡^ |  |
| Normal (f: 38.0-44.0; m: 39.5-50.5) | 216 | 36.9 | 110 | 53.1 | 106 | 28.0 |  | ⊥ (not ↓) |
| High (f: >44.0; m: >50.5) | 15 | 2.6 | 7 | 3.4 | 8 | 2.1 |  |  |
| *Inflammation* |  |  |  |  |  |  |  |  |
| C-reactive protein (mg/dL) |  |  |  |  |  |  |  |  |
| Median (IQR) | 586 | 43.62 (10.10 to 105.00) | 205 | 61.80 (25.45 to 129.50) | 381 | 33.60 (7.25 to 93.56) | <0.001^†^ | ↑ |
| Distribution – no./total no. (%) |  |  |  |  |  |  |  |  |
| Normal (≤0.5) | 8 | 1.4 | 1 | 0.5 | 7 | 1.8 | 0.179^‡^ | = (↑) |
| High (>0.5) | 578 | 98.6 | 204 | 99.5 | 374 | 98.2 |  |  |
| Distribution – no./total no. (%) |  |  |  |  |  |  |  |  |
| < 22 mg/dL | 205 | 35.0 | 44 | 21.5 | 161 | 42.3 | <0.001^†^ | ↑ (less ↓) |
| ≥ 22 mg/dL | 381 | 65.0 | 161 | 78.5 | 220 | 57.7 |  |  |
| Procalcitonin (ng/mL) |  |  |  |  |  |  |  |  |
| Median (IQR) | 130 | 0.13 (0.05 to 0.48) | 43 | 0.12 (0.05 to 0.28) | 87 | 0.16 (0.05 to 0.64) | 0.314^†^ | = |
| Distribution – no./total no. (%) |  |  |  |  |  |  |  |  |
| Normal (≤0.5) | 102 | 78.5 | 39 | 90.7 | 63 | 72.4 | 0.017^‡^ | ⊥ (not ↑) |
| High (>0.5) | 28 | 21.5 | 4 | 9.3 | 24 | 27.6 |  |  |

**Supplement Table 1 (cont.). Comparison of Standard Blood Laboratory Parameters between COVID-19 Positive and Negative tested Patients.**

| Parameters | N* | COVID-19  TOTAL | N* | COVID-19  Positive | N* | COVID-19  Negative | P value | Pattern |
| --- | --- | --- | --- | --- | --- | --- | --- | --- |
| *Blood chemistry* |  |  |  |  |  |  |  |  |
| Albumin (g/L) |  |  |  |  |  |  |  |  |
| Median (IQR) | 362 | 30.00 (26.00 to 36.00) | 55 | 30.00 (23.00 to 34.00) | 307 | 31.00 (26.00 to 36.00) | 0.207^†^ | = |
| Distribution – no./total no. (%) |  |  |  |  |  |  |  |  |
| Low (<35.0) | 252 | 69.6 | 44 | 80.0 | 208 | 67.8 | 0.080^§^ |  |
| Normal (35.0-52.0) | 110 | 30.4 | 11 | 20.0 | 99 | 32.2 |  | = (↓) |
| High (>52.0) | 0 | 0 | 0 | 0 | 0 | 0 |  |  |
| *Metabolism* |  |  |  |  |  |  |  |  |
| Glucose (mg/dL) |  |  |  |  |  |  |  |  |
| Median (IQR) | 541 | 112.00 (96.00 to 143.00) | 204 | 112.00 (99.25 to 141.50) | 337 | 112.00 (95.00 to 144.00) | 0.272^†^ | = |
| Distribution – no./total no. (%) |  |  |  |  |  |  |  |  |
| Low (<70) | 13 | 2.4 | 4 | 2.0 | 9 | 2.7 | 0.232^‡^ |  |
| Normal (70-100) | 151 | 27.9 | 49 | 24.0 | 102 | 30.3 |  | = (↑) |
| High (>100) | 377 | 69.7 | 151 | 74.0 | 226 | 67.0 |  |  |
| *Electrolytes* |  |  |  |  |  |  |  |  |
| Potassium (mmol/L) |  |  |  |  |  |  |  |  |
| Median (IQR) | 539 | 4.00 (3.70 to 4.20) | 183 | 4.00 (3.70 to 4.20) | 356 | 3.90 (3.60 to 4.20) | 0.149^†^ | = |
| Distribution – no./total no. (%) |  |  |  |  |  |  |  |  |
| Low (<3.5) | 65 | 12.1 | 17 | 9.3 | 48 | 13.5 | 0.285^†^ |  |
| Normal (3.5-5.5) | 469 | 87.0 | 165 | 90.2 | 304 | 85.4 |  | = (⊥) |
| High (>5.5) | 5 | 0.9 | 1 | 0.5 | 4 | 1.1 |  |  |
| Sodium (mmol/L) |  |  |  |  |  |  |  |  |
| Median (IQR) | 584 | 138.00 (135.00 to 140.00) | 207 | 136.00 (133.00 to 139.00) | 377 | 138.00 (136.00 to 140.00) | <0.001^†^ | ↓ |
| Distribution – no./total no. (%) |  |  |  |  |  |  |  |  |
| Low (<135) | 131 | 22.4 | 72 | 34.8 | 59 | 15.7 | <0.001^‡^ |  |
| Normal (135-150) | 449 | 76.9 | 133 | 64.2 | 316 | 83.8 |  | ⊥ (more ↓) |
| High (>150) | 4 | 0.7 | 2 | 1.0 | 2 | 0.5 |  |  |
| *Coagulation* |  |  |  |  |  |  |  |  |
| Activated partial thromboplastin time (sec) |  |  |  |  |  |  |  |  |
| Median (IQR) | 439 | 28.00 (25.00 to 32.50) | 141 | 29.90 (27.00 to 33.05) | 298 | 27.00 (24.00 to 31.00) | <0.001^†^ | ↑ |
| Distribution – no./total no. (%) |  |  |  |  |  |  |  |  |
| Low (<21) | 10 | 2.3 | 3 | 2.1 | 7 | 2.4 | <0.001^‡^ |  |
| Normal (21-32) | 316 | 72.0 | 85 | 60.3 | 231 | 77.5 |  | ⊥ (more ↑) |
| High (>32) | 113 | 25.7 | 53 | 37.6 | 60 | 20.1 |  |  |

**Supplement Table 1 (cont.). Comparison of Standard Blood Laboratory Parameters between COVID-19 Positive and Negative tested Patients.**

| Parameters | N* | COVID-19  TOTAL | N* | COVID-19  Positive | N* | COVID-19  Negative | P value | Pattern |
| --- | --- | --- | --- | --- | --- | --- | --- | --- |
| *Liver function* |  |  |  |  |  |  |  |  |
| Alanine aminotransferase (U/L) |  |  |  |  |  |  |  |  |
| Median (IQR) | 517 | 27.00 (17.00 to 50.00) | 171 | 32.00 (21.00 to 53.00) | 346 | 25.00 (15.00 to 45.00) | 0.001^†^ | ↑ |
| Distribution – no./total no. (%) |  |  |  |  |  |  |  |  |
| Normal (≤45) | 372 | 72.0 | 111 | 64.9 | 261 | 75.4 | 0.012^‡^ | ⊥ (more ↑) |
| High (>45) | 145 | 28.0 | 60 | 35.1 | 85 | 24.6 |  |  |
| Aspartate aminotransferase (U/L) |  |  |  |  |  |  |  |  |
| Median (IQR) | 329 | 33.00 (23.00 to 62.00) | 124 | 47.00 (29.00 to 70.00) | 205 | 26.00 (20.00 to 52.00) | <0.001^†^ | ↑ |
| Distribution – no./total no. (%) |  |  |  |  |  |  |  |  |
| Normal (≤35) | 176 | 53.5 | 46 | 37.1 | 130 | 63.4 | <0.001^‡^ | ↑ (not ↓) |
| High (>35) | 153 | 46.5 | 78 | 62.9 | 75 | 36.6 |  |  |
| Total Bilirubin (mg/dL) |  |  |  |  |  |  |  |  |
| Median (IQR) | 520 | 0.60 (0.40 to 0.80) | 173 | 0.60 (0.40 to 0.80) | 347 | 0.50 (0.40 to 0.90) | 0.311^†^ | = |
| Distribution – no./total no. (%) |  |  |  |  |  |  |  |  |
| Low (<0.3) | 47 | 9.0 | 11 | 6.3 | 36 | 10.4 | 0.005^‡^ |  |
| Normal (0.3-1.0) | 398 | 76.5 | 147 | 85.0 | 251 | 72.3 |  | ⊥ (less ↑) |
| High (>1.0) | 75 | 14.4 | 15 | 8.7 | 60 | 17.3 |  |  |
| Gamma glutamyl transpeptidase (U/L) |  |  |  |  |  |  |  |  |
| Median (IQR) | 517 | 43.00 (25.00 to 94.50) | 171 | 46.00 (27.00 to 88.00) | 346 | 41.00 (23.00 to 98.25) | 0.186^†^ | = |
| Distribution – no./total no. (%) |  |  |  |  |  |  |  |  |
| Normal (≤60) | 331 | 64.0 | 109 | 63.7 | 222 | 64.2 | 0.926^‡^ | = |
| High (>60) | 186 | 36.0 | 62 | 36.3 | 124 | 35.8 |  |  |
| Lipase (U/L) |  |  |  |  |  |  |  |  |
| Median (IQR) | 433 | 29.00 (18.00 to 50.50) | 151 | 39.00 (25.00 to 65.00) | 282 | 23.00 (14.00 to 41.00) | <0.001^†^ | ↑ |
| Distribution – no./total no. (%) |  |  |  |  |  |  |  |  |
| Normal (≤60.0) | 346 | 79.9 | 106 | 70.2 | 240 | 85.1 | <0.001^‡^ | ⊥ (more ↑) |
| High (>60.0) | 87 | 20.1 | 45 | 29.8 | 42 | 14.9 |  |  |

**Supplement Table 1 (cont.). Comparison of Standard Blood Laboratory Parameters between COVID-19 Positive and Negative tested Patients.**

| Parameters | N* | COVID-19  TOTAL | N* | COVID-19  Positive | N* | COVID-19  Negative | P value | Pattern |
| --- | --- | --- | --- | --- | --- | --- | --- | --- |
| *Heart function* |  |  |  |  |  |  |  |  |
| Creatine Kinase (U/L) |  |  |  |  |  |  |  |  |
| Median (IQR) | 517 | 111.00 (53.50 to 232.00) | 181 | 127.00 (63.00 to 243.50) | 336 | 102.50 (50.00 to 216.00) | 0.023^†^ | ↑ |
| Distribution – no./total no. (%) |  |  |  |  |  |  |  |  |
| Normal (≤190) | 359 | 69.4 | 113 | 62.4 | 246 | 73.2 | 0.011^‡^ | ⊥ (more ↑) |
| High (>190) | 158 | 30.6 | 68 | 37.4 | 90 | 26.8 |  |  |
| Lactate dehydrogenase (U/L) |  |  |  |  |  |  |  |  |
| Median (IQR) | 451 | 248.00 (199.00 to 331.00) | 157 | 285.00 (228.00 to 384.50) | 294 | 225.00 (189.00 to 297.75) | <0.001^†^ | ↑ |
| Distribution – no./total no. (%) |  |  |  |  |  |  |  |  |
| Normal (≤250) | 228 | 50.6 | 55 | 35.0 | 173 | 58.8 | <0.001^‡^ | ↑ (less ⊥) |
| High (>250) | 223 | 49.4 | 102 | 65.0 | 121 | 41.2 |  |  |
| *Renal function* |  |  |  |  |  |  |  |  |
| Creatinine (mg/dL) |  |  |  |  |  |  |  |  |
| Median (IQR) | 585 | 1.00 (0.80 to 1.30) | 208 | 1.00 (0.80 to 1.34) | 377 | 1.00 (0.80 to 1.30) | 0.081^†^ | = |
| Distribution – no./total no. (%) |  |  |  |  |  |  |  |  |
| Low (<0.5) | 12 | 2.1 | 0 | 0 | 12 | 3.2 | 0.016^‡^ |  |
| Normal (0.5-1.2) | 401 | 68.5 | 139 | 66.8 | 262 | 69.5 |  | ⊥ (less ↓) |
| High (>1.2) | 172 | 29.4 | 69 | 33.2 | 103 | 27.3 |  |  |
| Blood urea nitrogen (mg/dL) |  |  |  |  |  |  |  |  |
| Median (IQR) | 498 | 18.00 (12.00 to 28.00) | 178 | 17.50 (13.00 to 28.00) | 320 | 18.00 (12.00 to 28.00) | 0.555^†^ | = |
| Distribution – no./total no. (%) |  |  |  |  |  |  |  |  |
| Low (<6) | 10 | 2.0 | 1 | 0.6 | 9 | 2.8 | 0.227^‡^ |  |
| Normal (6-25) | 341 | 68.5 | 123 | 69.1 | 218 | 68.1 |  | = (⊥) |
| High (>25) | 147 | 29.5 | 54 | 30.3 | 93 | 29.1 |  |  |

Abbreviations: f, female; IQR, interquartile range; L, liter; m, male

* Number of parameters available.

^†^ Mann-Whitney U-test

^‡^ Chi-square test

^§^ Fisher’s exact test

**Supplement Table 2 Univariate Analyses and Area Under Receiver Operating Characteristic Curve of Comorbidities and Standard Blood Laboratory Parameters to distinguish between COVID-19 Positive and Negative tested Patients.**

| Parameters | OR (95% CI) | P value | AUC (95% CI) | P value |
| --- | --- | --- | --- | --- |
| *Comorbidities* |  |  |  |  |
| Chronic lung disease | 0.46 (0.28-0.77) | 0.003 | 0.451 (0.403-0.498) | 0.048 |
| Malignant tumor | 0.28 (0.15-0.54) | <0.001 | 0.440 (0.393-0.487) | 0.016 |
| Chronic liver disease | 0.15 (0.04-0.65) | 0.011 | 0.475 (0.427-0.523) | 0.310 |
| *Blood count* |  |  |  |  |
| Leukocytes (10^9^/L)  Leukocytes (ref: normal)  low  high  Leukocytes (ref: >10.0)  Leukopenia+normal (≤10.0) | 0.81 (0.77-0.86)  1.00  1.56 (0.83-2.92)  0.23 (0.15-0.36)  1.00  4.54 (2.90-7.11) | <0.001  0.168  <0.001  <0.001 | 0.277 (0.235-0.320)  0.349 (0.304-0.394)  0.640 (0.595-0.685) | <0.001  <0.001  <0.001 |
| Neutrophil-to-lymphocyte ratio  Neutrophil-to-lymphocyte ratio (ref: >2.33)  ≤2.33 | 1.00 (0.98-1.02)  1.00  2.97 (1.74-5.07) | 0.745  <0.001 | 0.443 (0.391-0.496)  0.561 (0.508-0.613) | 0.032  0.022 |
| Basophils 10^9^/L | 9.55*10^-18^  (6.02*10^-23^-1.51*10^-12^) | <0.001 | 0.300 (0.253-0.346) | <0.001 |
| Eosinophils (10^9^/L)  Eosinophils (ref: normal)  low  high  Eosinophils (ref: ≥0.1)  Eosinopenia (<0.1) | 0.00005 (0.000003-0.001)  1.00  5.72 (3.59-9.12)  0.93 (0.11-7.86)  1.00  5.74 (3.63-9.09) | <0.001  <0.001  0.946  <0.001 | 0.206 (0.164-0.249)  0.324 (0.278-0.370)  0.676 (0.630-0.722) | <0.001  <0.001  <0.001 |
| Monocytes (10^9^/L)  Monocytes (ref: normal)  low  high  Monocytes (ref: >0.9)  Monopenia+normal (≤0.9) | 0.26 (0.13-0.51)  1.00  0.00  0.43 (0.23-0.81)  1.00  2.30 (1.22-4.35) | <0.001  0.999  0.009  0.010 | 0.394 (0.344-0.445)  0.464 (0.414-0.515)  0.540 (0.489-0.590) | <0.001  0.177  0.134 |
| Thrombocytes (10^9^/L)  Thrombocytes (ref. >370)  Thrombopenia+normal (≤370) | 0.998 (0.997-1.00)  1.00  2.00 (1.10-3.66) | 0.013  0.024 | 0.423 (0.376-0.470)  0.531 (0.483-0.580) | 0.002  0.209 |
| Hemoglobin (g/dL)  Hemoglobin (ref: normal)  low  high  Hemoglobin (ref: f: <11.8; m: <13.5)  Normal+high (f: ≥11.8; m: ≥13.5) | 1.39 (1.27-1.52)  1.00  0.29 (0.20-0.42)  0.68 (0.11-4.14)  1.00  3.40 (2.38-4.86) | <0.001  <0.001  0.677  <0.001 | 0.697 (0.653-0.740)  0.647 (0.600-0.693)  0.648 (0.601-0.694) | <0.001  <0.001  <0.001 |
| *Inflammation* |  |  |  |  |
| C-reactive protein (U/L)  C-reactive protein (ref: <22 mg/dL)  ≥22 mg/dL | 1.004 (1.002-1.006)  1.00  2.68 (1.81-3.96) | <0.001  <0.001 | 0.610 (0.563-0.657)  0.604 (0.557-0.651) | <0.001  <0.001 |
| *Coagulation* |  |  |  |  |
| Activated partial thromboplastin time (sec)  Activated partial thromboplastin time  (ref: normal)  low  high  Activated partial thromboplastin time  (ref: <21sec)  Normal+high (≥21) | 1.04 (1.01-1.08)  1.00  1.17 (0.29-4.61)  2.40 (1.54-3.75)  1.00  1.12 (0.28-4.34) | 0.005  0.828  <0.001  0.885 | 0.619 (0.564-0.674)  0.586 (0.527-0.645)  0.501 (0.443-0.559) | <0.001  0.004  0.970 |

**Supplement Table 2 (continued) Univariate Analyses and Area Under Receiver Operating Characteristic Curve of Comorbidities and Standard Blood Laboratory Parameters to distinguish between COVID-19 Positive and Negative tested Patients.**

| Parameters | OR (95% CI) | P value | AUC (95% CI) | P value |
| --- | --- | --- | --- | --- |
| *Heart function* |  |  |  |  |
| Creatine Kinase (U/L)  Creatine kinase (ref: ≤190)  high (>190) | 1.00004 (0.9999-1.0002)  1.00  1.65 (1.12-2.42) | 0.675  0.011 | 0.561 (0.510-0.611)  0.554 (0.501-0.606) | 0.023  0.043 |
| Lactate dehydrogenase (U/L)  Lactate dehydrogenase (ref: ≤250)  high (>250) | 1.0005 (0.9996-1.001)  1.00  2.65 (1.77-3.96) | 0.274  <0.001 | 0.659 (0.607-0.711)  0.619 (0.565-0.673) | <0.001  <0.001 |
| *Liver function* |  |  |  |  |
| Alanine aminotransferase (U/L)  Alanine aminotransferase (ref: ≤45)  high (>45) | 0.999 (0.997-1.002)  1.00  1.66 (1.12-2.47) | 0.519  0.013 | 0.592 (0.541-0.642)  0.553 (0.499-0.606) | 0.001  0.051 |
| Aspartate aminotransferase (U/L)  Aspartate aminotransferase (ref: ≤35)  high (>35) | 1.000 (0.998-1.001)  1.00  2.94 (1.85-4.67) | 0.653  <0.001 | 0.657 (0.597-0.716)  0.632 (0.569-0.694) | <0.001  <0.001 |
| Lipase (U/L)  Lipase (ref: ≤60)  high (>60) | 1.002 (0.999-1.005)  2.43 (1.50-3.92) | 0.137  <0.001 | 0.684 (0.632-0.735)  0.575 (0.517-0.632) | <0.001  0.011 |
| *Renal function* |  |  |  |  |
| Creatinine (ref. normal)  Low  High | 1.00  0.00  1.26 (0.87-1.82) | 0.999  0.214 | 0.540 (0.491-0.588) | 0.110 |

Abbreviation: AUC, area under the curve; CI, confidence interval; OR, odds ratio.
